# Altered microRNA expression in severe COVID-19: potential prognostic and pathophysiological role

**DOI:** 10.1101/2022.03.21.22272480

**Authors:** Nathalie Garnier, Kato Pollet, Marie Fourcot, Morgan Caplan, Guillemette Marot, Julien Goutay, Julien Labreuche, Fabrice Soncin, Rabah Boukherroub, Didier Hober, Lille COVID Research Network (LICORNE), Sabine Szunerits, Julien Poissy, Ilka Engelmann

## Abstract

**Background:** The severe acute respiratory syndrome coronavirus 2 (SARS-CoV-2) pandemic is ongoing. The pathophysiology of SARS-CoV-2 infection is beginning to be elucidated but the role of microRNAs (miRNAs), small non-coding RNAs that regulate gene expression, remains incompletely understood. They play a role in the pathophysiology of viral infections with potential use as biomarkers. The objective of this study was to identify miRNAs as biomarkers of severe COVID-19 and to analyze their role in the pathophysiology of SARS-CoV-2 infection.

**Methods:** miRNA expression was measured in nasopharyngeal swabs from 20 patients with severe COVID-19, 21 patients with non-severe COVID-19 and 20 controls. Promising miRNAs to differentiate non-severe from severe COVID-19 patients were identified by differential expression analysis and sparse Partial Least Squares-Discriminant Analysis (sPLS-DA). ROC analysis, target prediction, GO enrichment and pathway analysis were used to analyze the role and the pertinence of these miRNAs in severe COVID-19.

**Results:** The number of expressed miRNAs was lower in severe COVID-19 patients compared to non-severe COVID-19 patients and controls. Among the differentially expressed miRNAs between severe COVID-19 and controls, 5 miRNAs were also differentially expressed between severe and non-severe COVID-19. sPLS-DA analysis highlighted 8 miRNAs, that allowed to discriminate the severe and non-severe COVID-19 cases. Target and functional analysis revealed enrichment for genes involved in viral infections and the cellular response to infection as well as one miRNA, hsa-miR-15b-5p, that targeted the SARS-CoV-2 RNA.

The comparison of results of differential expression analysis and discriminant analysis revealed three miRNAs, namely hsa-miR-125a-5p, hsa-miR-491-5p and hsa-miR-200b-3p. These discriminated severe from non-severe cases with areas under the curve ranging from 0.76 to 0.80.

**Conclusions:** Our analysis of miRNA expression in nasopharyngeal swabs revealed several miRNAs of interest to discriminate severe and non-severe COVID-19. These miRNAs represent promising biomarkers and possibly targets for antiviral or anti-inflammatory treatment strategies.

## Background

At the end of December 2019, an outbreak caused by a novel coronavirus, later named severe acute respiratory syndrome coronavirus 2 (SARS-CoV-2), was announced in Wuhan, China [1, 2]. *Coronaviridae* are enveloped viruses with a single-stranded positive-sense RNA genome of approximately 30 kb [3]. The *Coronaviridae* family comprises seven members that cause disease in humans, namely HCoV-229E, HCoV-OC43, HCoV-NL63, HCoV-HKU1, SARS-CoV, SARS-CoV-2 and MERS-CoV [2, 3]. SARS-CoV-2 rapidly spread outside of China and caused a pandemic [4]. The disease caused by SARS-CoV-2 was named coronavirus disease 2019 (COVID-19). Clinical manifestations range from asymptomatic infection to severe respiratory failure and multiorgan dysfunction. Around 15% of infected patients will need hospitalization, and 5% will required ICU management. An imbalanced host response, characterized by reduced type I interferon (IFN-I; IFN-III) response to SARS-CoV-2 in combination with elevated chemokines and high expression of interleukin 6 (IL-6) has been suggested to drive the development of severe and critical COVID-19 [5]. Management of these severely ill patients has been improved by the introduction of corticosteroids since the Recovery trial [6]. Anti-IL-6 drugs and JAK-inhibitors are also recommended [7] but some debate persists regarding patients non represented in clinical trials and on the fact to use these drugs systematically with corticosteroids or after evaluation of response to corticoids [8]. The best way to manage the respiratory failure is also a matter of debate, non-invasive procedures being an option [9–11], but with the risk to delay intubation in case of worsening [12].

In this context, we need to be able to stratify patients according to their risk to develop severe COVID-19 in order to be able to adapt therapeutic strategy in a “personalized approach”. Interleukin-6 (IL-6) [13] and SARS-CoV-2 RNAemia were described to be associated with disease severity [14, 15], but are not really helpful at bedside for the early identification of patients at risk of worsening, in order to adapt the therapeutic strategy.

MicroRNAs (miRNAs) are a class of small, non-coding RNAs with an average length of 22 nucleotides that regulate gene expression on the posttranscriptional level [16]. miRNAs are involved in virtually all physiological and pathological processes including viral infections and the antiviral immune response [17]. Studies investigating the role of miRNAs in infections caused by different members of the *Coronaviridae* family have shed some light on the pathophysiology of *Coronaviridae* infections [18–23]. miRNAs also have the potential to be highly specific biomarkers for diagnosis of viral infections [24] as well as biomarkers for disease severity [25, 26].

The aims of the current study were (1) to identify human miRNAs that can be used as biomarkers for severe disease and (2) unravel the potential role of these miRNAs in the pathophysiology of SARS-CoV-2 infection.

## Methods

### Patients and specimens

A total of 61 patients were included in the study, including 20 patients with severe COVID-19, 21 patients with non-severe COVID-19 and 20 controls (Supplementary table 1). Severe COVID-19 patients were patients admitted to the intensive care unit for oxygen treatment. Non-severe COVID-19 patients were seen in ambulatory medicine and not hospitalized, without requirement of oxygen treatment. The control group consisted of 20 patients that had nasopharyngeal swab specimens performed for routine diagnostic purposes and who did not have COVID-19. Nasopharyngeal swab specimens were obtained from all patients for routine diagnostic purposes by using flocked swabs that were placed in universal transport medium. Specimens were stored at −80°C. The first available specimen was used and median time of sampling was 7 days after symptom onset (Supplementary Table 1). Demographic data were retrospectively collected from hospital charts and the virology laboratory database. The study was approved by the French Institutional Authority for Personal Data Protection (Commission Nationale de l’Informatique et des Libertés DR-2020-178, October 22nd, 2020) and the ethics committee (Comité de Protection des Personnes Nord Ouest IV, ECH20/09, September 7th, 2020).

### RNA extraction

RNA extraction was performed by using the MagMAX mirVana Total RNA Isolation Kit (Thermofisher Scientific, Courtaboeuf, France) according to the manufacturer’s instructions.

### miRNA profiling

Poly-A-tailing, adapter ligation, reverse transcription and pre-amplification was performed using the TaqMan Advanced miRNA cDNA Synthesis Kit (Thermofisher Scientific, Courtaboeuf, France) according to the manufacturer’s instructions. Quantitative PCR was done with TaqMan Fast Advanced Master Mix and the TaqMan™ Advanced miRNA Human A and B Cards (Thermofisher Scientific, Courtaboeuf, France) on a QuantStudio 7 Flex instrument (Thermofisher Scientific, Courtaboeuf, France) according to the manufacturer’s instructions. The expression of miRNAs was quantified using the online application of the Thermofisher Cloud (www.thermofisher.com). Amplification data were reviewed and the thresholds adapted.

In the following of this paper a miRNA is called “detected” if its Ct-value was strictly lower than 40. We considered as “expressed” a miRNA detected in at least 80% of the nasopharyngeal swab specimens of at least one of the three groups. Only the expressed miRNAs were kept for further analyses

### Target prediction, GO enrichment and pathway analysis

miRWalk (http://mirwalk.umm.uni-heidelberg.de/) was used for the prediction of targets of dysregulated miRNAs and GO enrichment analysis, Kegg and Reactome pathways enrichment analyses [27]. Only validated target genes were considered for GO enrichment analysis, Kegg and Reactome pathways. An adjusted p-value <0.05 was considered statistically significant. miRDB was used for the prediction of miRNAs that target the SARS-CoV-2 genome (NC_045512.2) [28]. Only miRNAs with a target score of at least 90 were considered.

### Statistical analysis

Statistical analysis was performed by using R (version 4.1.2). Following the idea of global mean normalization [29] but reproducing the Thermofisher cloud normalization, the data were normalized by subtracting from each Ct value the median Ct value of the plate from which it originated. The delta delta Ct method was used to calculate fold changes of miRNA expression between groups [30].

To compare the number of expressed miRNAs between groups, a Kruskal-Wallis test was performed followed by Dunn’s post-hoc test. Then differential expression analysis of miRNA between the severe COVID-19 group and the control group were performed using Mann-Whitney U tests. Raw p-values of these multiple univariate analyses were adjusted with Benjamini-Hochberg method [31], which controls the False Discovery Rate. miRNAs presenting an adjusted p-value of <0.05 were considered differentially expressed between the severe COVID-19 group and the control group. Expression of these differentially expressed miRNAs was then compared between severe and non-severe COVID-19 groups by using Mann-Whitney U tests and Bonferroni correction to control the Family-Wise Error Rate (Probability of having at least one false positive). An adjusted p-value of <0.05 was considered statistically significant.

In parallel to these univariate analyses sparse Partial Least Squares-Discriminant Analysis (sPLS-DA) was performed with the mixOmics package [32]. This multivariate analysis enables to discriminate severe and non-severe COVID-19 groups. It is appropriate to this dataset which contains much more variables than individuals as PLS can deal with correlated variables and sparsity ensures variable selection. The number of components was set to 2 in order to facilitate visualization.

Linear discriminant analyses were run with the lda function of the MASS R package on the subset of miRNAs selected by sPLS-DA. Leave-one-out cross validation was used to predict the severity of each Covid-19 patient from these 8 miRNAs. A confusion matrix was built to compare the prediction with the true group and evaluate the potential of prediction of these candidate biomarkers.

Receiver operating characteristic (ROC) analysis with area under the curve (AUC) was performed to analyze the diagnostic performance to differentiate severe and non-severe COVID-19 of promising miRNAs defined by the consensus between differential expression analysis and sPLS-DA.

## Results

### miRNA expression in severe COVID-19 patients compared to controls

The expression of 754 miRNAs was measured in 61 nasopharyngeal swab specimens from patients with severe COVID-19 (n=20), non-severe COVID-19 (n=21) and patients without SARS-CoV-2 infection (controls, n=20) (Supplementary Table 1). 194 (25.7%) miRNAs were detected in at least 80% of the nasopharyngeal swab specimens of at least one of the groups and were considered as expressed for further analyses. The number of expressed miRNAs differed between the severe group (median 151 miRNAs) and the other groups (median 184 for the non-severe and 190.5 for the control groups, respectively) (Figure 1), with the severe COVID-19 group having less miRNAs detected than the other two groups. Next, quantitative expression of miRNAs was analyzed. 14 miRNAs were differentially expressed in severe COVID-19 versus controls (p < 0.05, Table 1). Interestingly, all 14 miRNAs showed lower expression in the severe COVID-19 group compared to the control group.

**Table 1.** miRNAs differentially expressed between severe COVID-19 and controls and non-severe COVID-19.

| miRNA | Log<sub>2</sub> fold-change |  | Log<sub>2</sub> fold-change |  |
| --- | --- | --- | --- | --- |
|  | severe COVID-19<br>versus control | adjusted p-value | severe COVID-19<br>versus non-severe<br>COVID-19 | adjusted p-value |
| hsa-miR-125a-5p | -5.170 | 0.044 | -4.853 | 0.045 |
| hsa-miR-200b-3p | -3.012 | 0.044 | -2.752 | 0.013 |
| hsa-miR-200c-3p | -2.328 | 0.044 | -1.280 | NS |
| hsa-miR-218-5p | -6.534 | 0.044 | -5.165 | NS |
| hsa-miR-27a-3p | -2.848 | 0.044 | -1.790 | NS |
| hsa-miR-30c-5p | -8.041 | 0.044 | -7.232 | NS |
| hsa-miR-30d-5p | -3.327 | 0.044 | -1.717 | NS |
| hsa-miR-375 | -1.977 | 0.044 | -0.859 | NS |
| hsa-miR-378a-3p | -4.365 | <0.001 | -1.919 | NS |
| hsa-miR-422a | -5.682 | 0.002 | 0.067 | NS |
| hsa-miR-455-5p | -2.724 | 0.044 | -2.260 | 0.034 |
| hsa-miR-532-5p | -1.898 | 0.044 | -0.499 | NS |
| hsa-miR-340-5p | -5.601 | 0.044 | -4.744 | 0.011 |
| hsa-miR-491-5p | -4.887 | 0.044 | -4.326 | 0.031 |
| NS : not significant |  |  |  |  |

**Figure 1.**
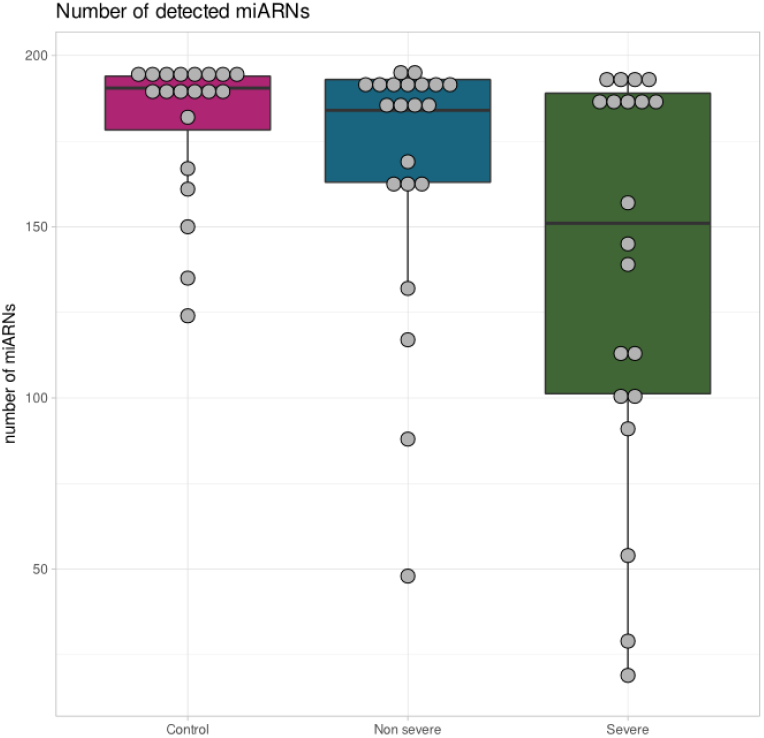
Reduced miRNA expression in severe COVID-19. Number of miRNAs detected in the severe, non-severe COVID-19 and controls. Box plots showing median and interquartile ranges of the number of expressed miRNAs in each group.

### miRNA expression in severe COVID-19 compared to non-severe COVID-19

Five of the miRNAs that were differentially expressed between the severe COVID-19 and the control group, namely hsa-miR-125a-5p, hsa-miR-200b-3p, hsa-miR-340-5p, hsa-miR-455-5p and hsa-miR-491-5p, were also downregulated in the severe versus the non-severe COVID-19 groups (Figure 2 A,B; Table 1). They showed −2.3 to −4.9 log_2_ fold changes in the severe compared to the non-severe COVID-19 group (Table 1).

**Figure 2.**
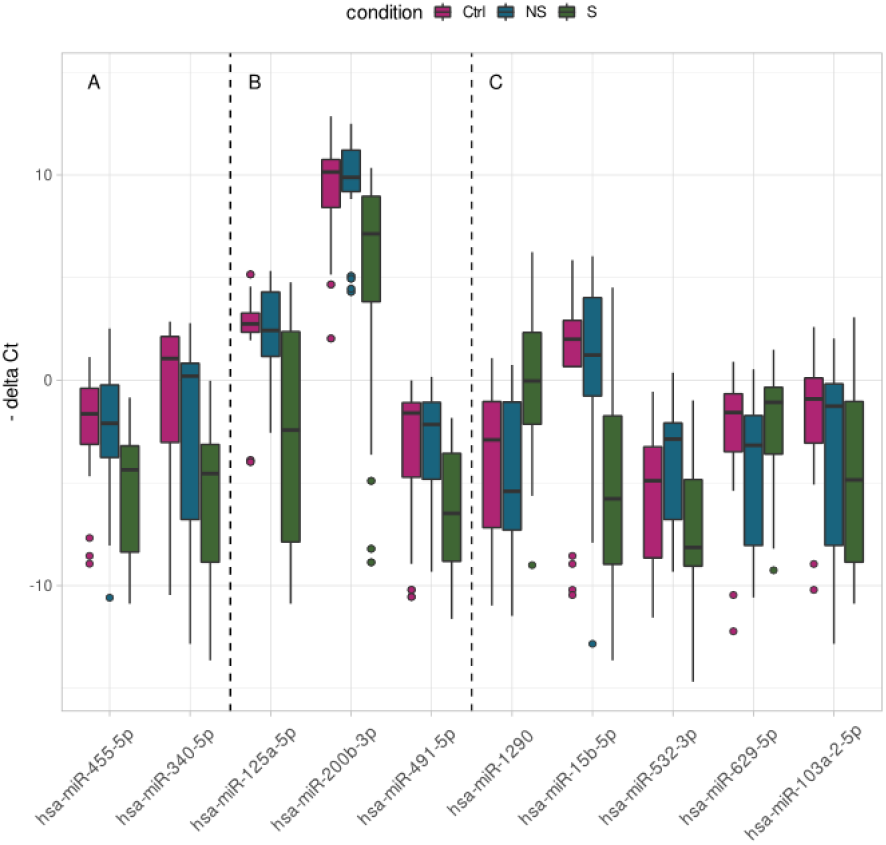
miRNA expression discriminates severe and non-severe COVID-19. Box-plot showing the expression of miRNAs that are differentially expressed between severe (S) and non-severe (NS) COVID-19 and control groups (Ctrl) in univariate analysis (A,B). Box-plot showing the expression of miRNAs that discriminate severe and non-severe COVID-19 by sPLS-DA (B,C). The miRNAs identified by both analyses are shown in B. The bars show the median and the boxes the interquartile ranges.

In parallel, sPLS-DA multivariate analysis was performed (Figure 3). This analysis selected 8 miRNAs: hsa-miR-125a-5p, hsa-miR-1290, hsa-miR-15b-5p, hsa-miR-491-5p, hsa-miR-532-3p, hsa-miR-200b-3p, hsa-miR-629-5p, hsa-miR-103a-2-5p, that discriminated severe and the non-severe COVID-19 (Figure 2 B,C). Six of these miRNAs were downregulated whereas two were upregulated in severe COVID-19 (Figure 2 B,C).

**Figure 3.**
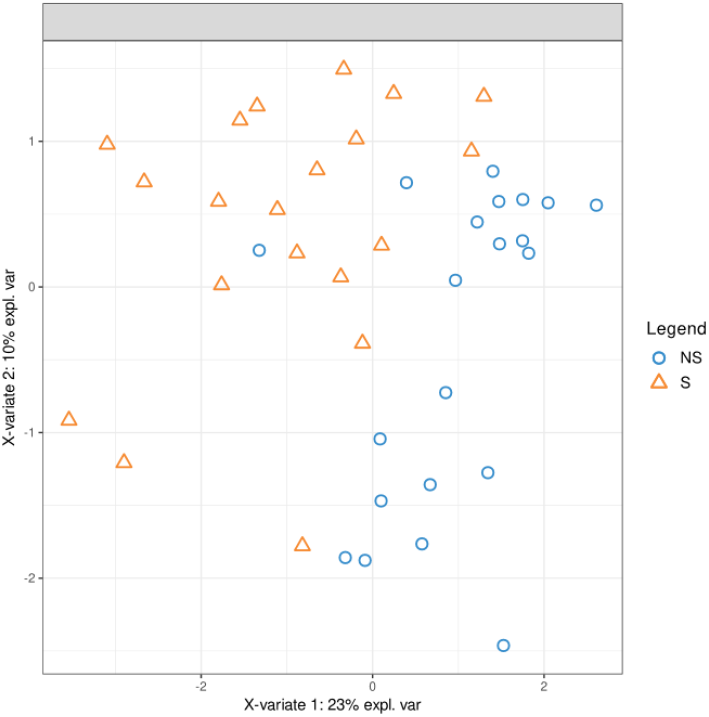
miRNA expression discriminates severe and non-severe COVID-19. Severe (S, triangles) and non-severe COVID-19 cases (NS, circles) are discriminated by miRNA expression by sPLS-DA analysis.

5 out of 8 of the miRNAs selected by sPLS-DA were not differentially expressed in univariate analyses (Figure 2C) but are necessary to build scores with linear discriminant analysis. When using leave-one-out cross validation, 35 out of 41 patients (85.37%) were correctly clustered (Table 2).

**Table 2.** Confusion matrix for prediction of Covid19 severity from miRNA expression with linear discriminant analysis.

|  | Predicted as non-severe COVID-19 | Predicted as severe COVID-19 |
| --- | --- | --- |
| non-severe COVID-19 (n=21) | 19 | 2 |
| severe COVID-19 (n=20) | 4 | 16 |

The comparison of results of differential expression analysis and sPLS-DA revealed three miRNA in common that were of particular interest, namely hsa-miR-125a-5p, hsa-miR-491-5p and hsa-miR-200b-3p (Figure 2B). These miRNAs discriminated severe from non-severe cases with areas under the curve (AUC) ranging from 0.76 to 0.79 (Figure 4).

**Figure 4.**
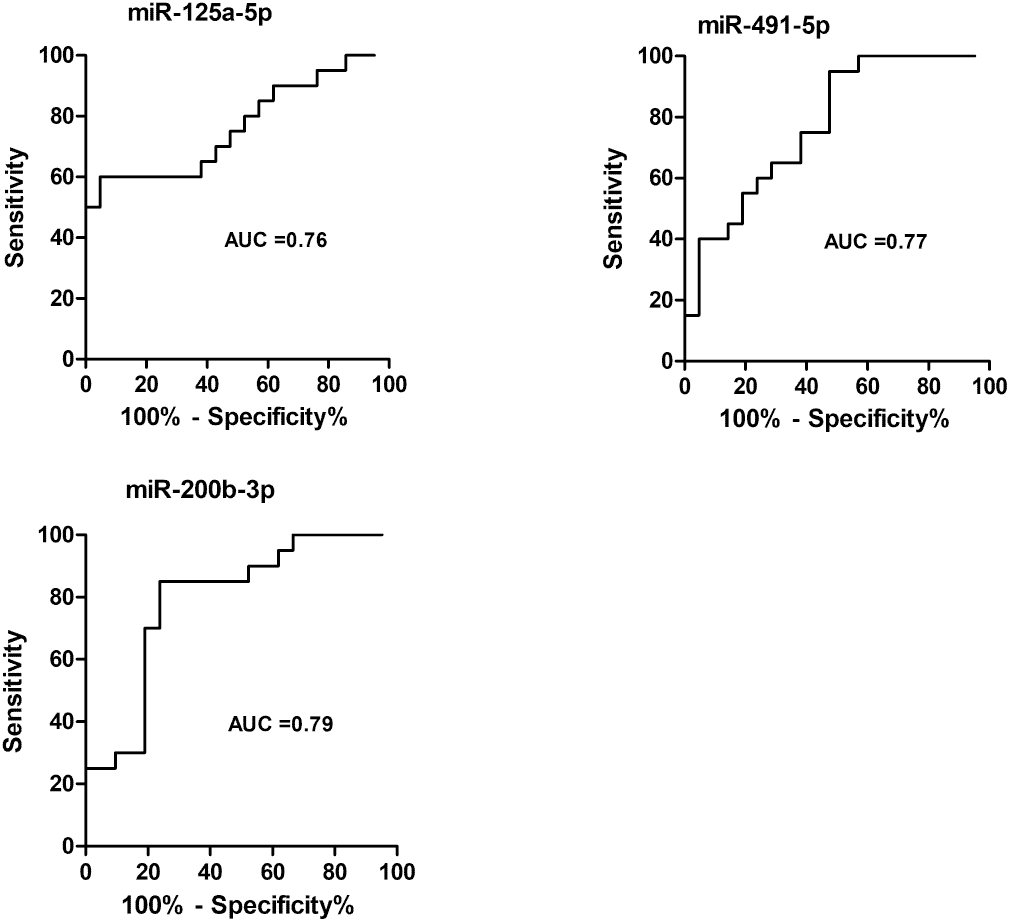
ROC analysis for the discrimination of severe and non-severe COVID-19. ROC curve and AUC values for the classification into severe and non-severe COVID-19 based on miRNA expression are shown for the three miRNAs identified in common by sPLS-DA and univariate analysis.

### Role of miRNAs in the pathophysiology of severe COVID-19

We hypothesized that the 10 miRNAs selected by sPLS-DA and/or differential analysis (Figure 2 A,B,C) play a role in the pathophysiology of severe COVID-19. To analyze their role, target prediction was performed using miRWalk [27]. 95 validated target genes were retrieved (Supplementary table 2). GO enrichment analysis was performed to analyze the function of these genes. Five, eight and zero GO terms associated to molecular function, biological process and cellular component were enriched in the target genes, respectively (Supplementary Tables 3 and 4). Enriched biological processes were involved in DNA damage, ubiquitination and antigen processing and presentation (Supplementary Table 3). Enriched molecular functions were involved in protein kinase activity, ubiquitination and RNA polymerase II activity (Supplementary Table 4). Kegg pathway analysis revealed enrichment in pathways that play a role in ubiquitination, viral infections and the immune response (Table 3). Reactome pathway analysis showed enrichment for pathways involved in antigen processing, NFkappaB and other signaling pathways (Table 4).

**Table 3.** Kegg pathways enrichment analysis.

| Kegg pathway | p-value | adjusted p-value (BH) |
| --- | --- | --- |
| hsa04120_Ubiquitin_mediated_proteolysis | 0.001 | 0.004 |
| hsa05162_Measles | 0.001 | 0.004 |
| hsa05169_Epstein-Barr_virus_infection | 0.001 | 0.004 |
| hsa05170_Human_immunodeficiency_virus_1_infection | 0.002 | 0.004 |
| hsa04218_Cellular_senescence | 0.002 | 0.004 |
| hsa04630_JAK-STAT_signaling_pathway | 0.003 | 0.004 |
| hsa05166_Human_T-cell_leukemia_virus_1_infection | 0.008 | 0.012 |
| hsa05131_Shigellosis | 0.011 | 0.014 |
| hsa05200_Pathways_in_cancer | 0.032 | 0.035 |

**Table 4.** Reactome pathway enrichment analysis.

| Reactome pathway | p-value | adjusted p-value (BH) |
| --- | --- | --- |
| R-HSA-983168_Antigen processing: Ubiquitination & Proteasome degradation | <0.001 | <0.001 |
| R-HSA-2871837_FCERI mediated NF-kB activation | <0.001 | <0.001 |
| R-HSA-5607764_CLEC7A (Dectin-1) signaling | <0.001 | <0.001 |
| R-HSA-9020702_Interleukin-1 signaling | <0.001 | <0.001 |
| R-HSA-202424_Downstream TCR signaling | <0.001 | <0.001 |

### miRNAs targeting SARS-CoV-2

We next searched for miRNAs that may target the SARS-CoV-2 genome among the miRNAs associated with severe COVID-19. We found that hsa-miR-15b-5p could target the SARS-CoV-2 genome according to miRDB [28], with a score of 99 and 16 predicted miRNA seed binding positions in the SARS-CoV-2 genome (Table 5). hsa-miR-15b-5p was downregulated in severe COVID-19 in our study (Figure 2C).

**Table 5.**
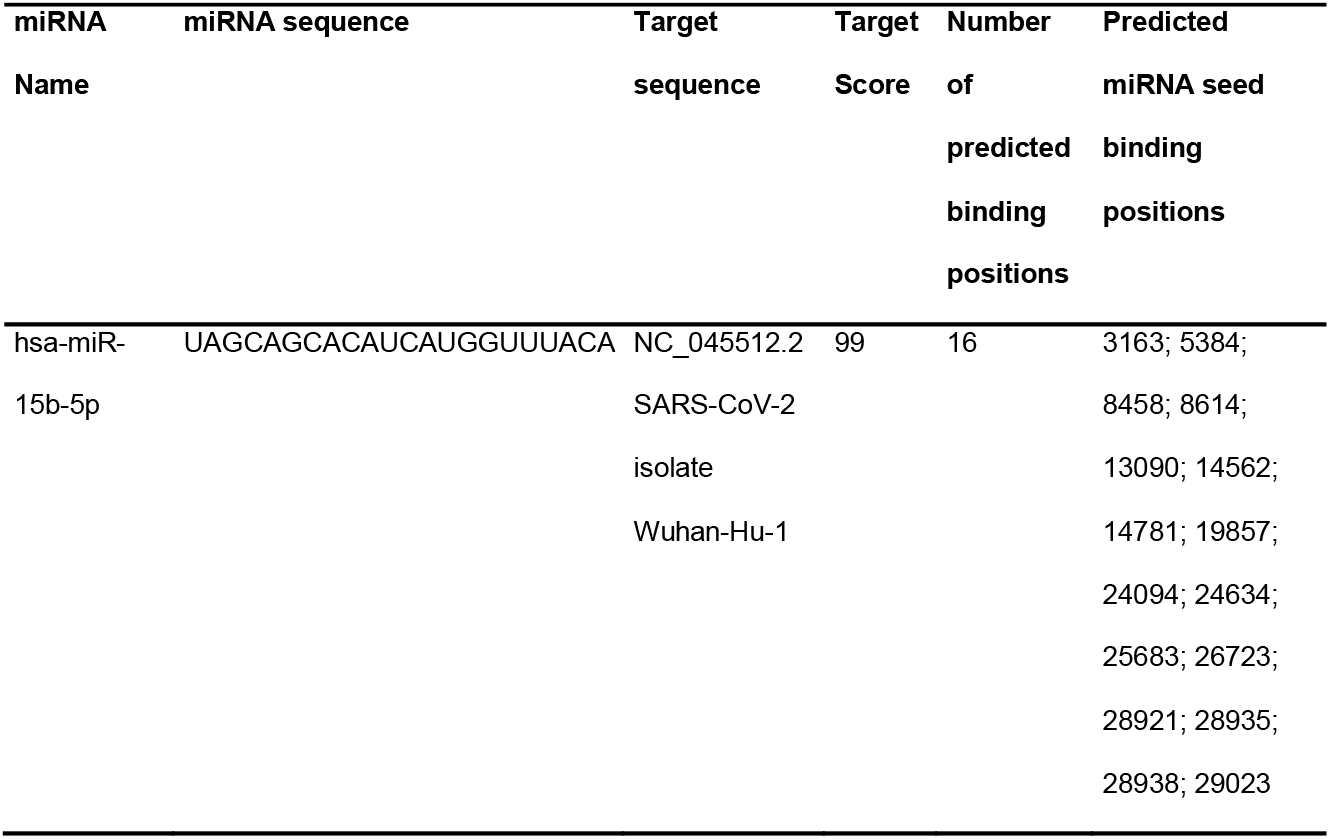
miRDB prediction of miRNAs binding to SARS-CoV-2 genome.

## Discussion

The role of miRNAs in SARS-CoV-2 infection is incompletely understood. Some recent studies have started to shed light on the involvement of miRNAs in SARS-CoV-2 infection, with most of these studies being based on computational prediction and *in silico* analyses [33]. A few experimental studies on the expression of miRNAs in patients with COVID-19 have been performed [34–45]. Whereas these studies have investigated miRNA expression in the blood, our study and a recently published study by Eichmeier and colleagues [46] are the first to investigate the miRNA expression profile in nasopharyngeal swabs of COVID-19 patients. Eichmeier and colleagues focused on the differential expression of miRNAs between SARS-CoV-2 RNA positive and negative nasopharyngeal specimens but no details were given on the patients [46]. We chose to investigate miRNA expression in nasopharyngeal swabs because we reasoned that (i) it would be useful to identify a biomarker of disease severity in the same specimen type used for diagnostics of COVID-19 and (ii) miRNA expression in nasopharyngeal swabs may reveal insights into pathophysiology of the disease.

Concerning the role of miRNAs in the physiopathology of COVID-19, the first important finding of our study is that the number of miRNAs expressed in nasopharyngeal secretions of severe COVID-19 patients was lower than in non-severe COVID-19 and controls. Secondly, most of the miRNAs associated with severe COVID-19 were found to be downregulated. This is in agreement with a recent study where a paralleled downregulation of miRNA expression in severe COVID-19 was reported [47]. Bartoszewski et al. suggested that the SARS-CoV-2 genome acts as miRNA sponge to reduce cellular miRNA levels [48]. In plants and insects, RNA silencing pathways have been identified as antiviral defense mechanisms and recent findings suggest that they also have antiviral functions in mammalian cells [49]. To escape this antiviral defense, plant and insect viruses possess virus-encoded suppressors of RNA interference [50]. There is some evidence that viruses also interfere with miRNA biogenesis in mammals [50–52]. An interesting hypothesis to explain our findings is thus that SARS-CoV-2 might directly target cellular miRNA biogenesis. A finding that supports this hypothesis is that the mRNA expression level of essential components of the RNA interference machinery Ago2, Dicer and Drosha were found significantly downregulated in COVID-19 patients [53]. Furthermore, miRNA depletion enhances proinflammatory cytokine production, including expression of IL-6 [54]. Interestingly, the global repression of miRNA expression in severe COVID-19 that we observed in the current study may thus be causally linked to the hyperinflammatory state found in severe COVID-19.

Next, we searched for specific miRNAs that were associated with severe COVID-19. Univariate analyses revealed that five miRNAs (hsa-miR-125a-5p, hsa-miR-200b-3p, hsa-miR-340-5p, hsa-miR-455-5p and hsa-miR-491-5p) were differentially expressed between patients with severe COVID-19 and patients with non-severe COVID-19 and controls (Figure 2 A,B). Interestingly, hsa-miR-455-5p was also found downregulated in the plasma of COVID-19 patients compared to controls [39] and the hsa-miR-125a-5p/miR-133a-3p ratio differed between critically ill COVID-19 and non-COVID-19 patients in bronchial aspirate specimens [55]. Multivariate sPLS-DA analysis highlighted 8 miRNAs discriminating between severe and non-severe COVID-19 (Figure 2B,C). The high percentage of correctly predicted severity by using the expression of these miRNAs (Table 2) suggests that these candidates could be used in scores to cluster patients. However, the coefficients of these scores should be built from a future study that includes more patients and validated in an independent cohort. There were three miRNAs in common with the univariates analyses and the multivariate analysis, namely hsa-miR-125a-5p, hsa-miR-491-5p and hsa-miR-200b-3p. These miRNAs are candidate biomarkers for severe COVID-19 according to ROC analysis with AUCs up to 0.79 (Figure 4). In future research and to evaluate their performance to predict disease severity, it would be interesting to determine their expression at different time points, i.e. before, during and after resolution of severe COVID-19.

In total we found ten miRNAs of interest to discriminate between severe and non-severe COVID-19 (Figure 2 A,B,C). We hypothesized that these miRNAs are implicated in the pathophysiology of severe COVID-19 and we analyzed the function of confirmed target genes of these miRNAs. Taken together, the results showed that the target genes were involved in several processes known to be implicated in viral infections, the cellular response to infection, ubiquitination and antigen processing and presentation (Tables 3 and 4, Supplementary Tables 2,3,4).

Of the miRNAs found of interest to discriminate between severe and non-severe COVID-19 in our study, of note, some were reported to be implicated in viral and other infections: Interestingly, hsa-miR-125a-5p and miRNAs from miR-200 family were recently reported to target the 3’ UTR of ACE2 mRNA [56]. A decrease in their expression as observed in our study would therefore result in upregulation of ACE2, the receptor of SARS-CoV-2. Furthermore, hsa-miR-125a-5p was reported to interact with the hepatitis B virus (HBV) genome and interfered with HBV replication and translation *in vitro* [57, 58]. hsa-miR-125a-5p levels in liver biopsies of hepatitis B infected patients correlated with liver and plasma HBV-DNA values and were predictive of disease progression [59], and the authors suggested that this miRNA may be part of a negative feedback limiting HBV replication.

hsa-miR-1290 was upregulated in A549 cells after influenza virus infection [60]. Plasma hsa-miR-1290 expression enabled differentiation of necrotizing enterocolitis cases from neonatal sepsis cases [61]. miR-1290 showed higher expression in latently HIV-1 infected cells than in productively infected cells. This miRNA targeted the 3’ untranslated region of HIV-1 and reduced its replication and is hypothesized to play a role in HIV-1 latency [62].

Furthermore, levels of hsa-miR-200b-3p were lower in human cytomegalovirus (HCMV)-infected gastro-intestinal tissues and negatively correlated with HCMV levels [63]. hsa-miR-200b-3p targeted a HCMV protein, repressed replication and was reported to play a role in HCMV latency [64]. Higher levels of hsa-miR-200b-3p were associated with less HCMV replication after solid organ transplantation [65].

Both hsa-miR-200b and hsa-miR-455 were found upregulated in pneumonia [66]. hsa-miR-455-5p was overexpressed in patients with neonatal sepsis compared to healthy controls and highly expressed miR-455-5p was associated with poor prognosis [67]. miR-455-5p was upregulated in rabies virus infection *in vitro*, decreased SOCS3 expression and increased STAT3 activity resulting in enhanced viral replication and the production of interleukin-6 (IL-6) [68]. Furthermore, this miRNA targeted the C-C motif chemokine receptor 5 (CCR5) [69]. CCR5 is a chemokine receptor involved in host responses and host susceptibility, especially to viruses, and is highly expressed in lung tissue [70]. Of interest, CCR5 is involved in severe COVID-19 and has been proposed as anti-inflammatory treatment target [70–72]. hsa-miR-455-5p was reported to have an anti-inflammatory role in multiple sclerosis [73]. Taken together, this underlines the role of hsa-miR-455-5p in viral infections and the inflammatory response and its potential as target of therapeutic interventions.

hsa-miR-340-5p was downregulated in A549 cells infected with influenza A virus or other RNA viruses. Its overexpression enhanced influenza A virus replication whereas its inhibition decreased influenza A virus replication [74]. hsa-miR-340-5p reduced the expression of CCL5, CXCL10, IL-6, and IFNβ in influenza A virus infected cells. RIG-I and OAS2 were identified as targets of hsa-miR-340-5p [74]. Interestingly, this miRNA was downregulated in the blood of COVID-19 patients [34] and in our study, suggesting it may be involved in the pathophysiology of COVID-19.

hsa-miR-532-3p diminished the levels of ASK1 and downstream phosphorylation/translocation of p38 MAPK during LPS/TNFα induced inflammation in macrophages and reduced the release of various pro-inflammatory cytokines and chemokines, including IL-6 and TNF-alpha [75]. IL-6 is a proinflammatory cytokine that has been reported to be involved in the cytokine storm observed in severe COVID-19 [13]. Consequently, IL-6 inhibition has received much attention as potential treatment. Especially tocilizumab, a recombinant humanized monoclonal antibody directed against the interleukin-6 (IL-6) receptor is under investigation for the treatment of COVID-19 and has been shown to decrease mortality in the Recovery trial and in the WHO meta-analysis [76–80]. The downregulation of anti-inflammatory miRNAs, such as hsa-miR-532-3p, hsa-miR-340-5p and hsa-miR-455-5p, in severe COVID-19 (Table 1) is in line with a hyperinflammatory state in severe COVID-19. Supplementation of these anti-inflammatory miRNAs may represent a novel therapeutic strategy.

hsa-miR-15b-5p was downregulated in severe COVID-19 in our study (Figure 2C). Interestingly, hsa-miR-15b-5p was also found downregulated in the lungs of hamsters infected with SARS-CoV-2 [81] and dysregulated in the blood of severe COVID-19 patients [37]. This miRNA was predicted to bind to the SARS-CoV-2 genome with a high confidence score in our study and also in another recent study [82]. Furthermore, hsa-miR-15b-5p interacted with the RNA component of viral RdRp and S protein *in vitro* [83, 84]. Taken together, this suggests that the downregulation of hsa-miR-15b-5p may represent a mechanism of SARS-CoV-2 to escape the host antiviral defense.

### Study limitations

our study has a relatively small size. Therefore our results should be confirmed in a larger cohort, which could allow a multiparametric approach and the building of scores. Another limit is the heterogeneity of the control group with respect to clinical severity. Moreover, our study showed several miRNAs allowed to differentiate between severe and non-severe COVID-19. In order to be used as predictive biomarkers, the expression of these miRNAs should be studied at disease onset and during the clinical course in order to determine the kinetics of miRNA expression and their correlation with severity.

## Conclusions

Taken together, we found that the number of expressed miRNAs was lower in in severe COVID-19 patients compared to non-severe COVID-19 patients and controls. Several miRNAs were associated with severe COVID-19 and functional analysis of these miRNAs suggested a role in the pathophysiology of the disease. Further characterization of their implication in SARS-CoV-2 infection will enable elucidation of the molecular mechanisms and may reveal potential targets for antiviral or anti-inflammatory treatment of COVID-19.

## Data Availability

The data that support the findings of this study are available from the corresponding author upon reasonable request.

## Acknowledgements

The authors thank B. Guerred, S. Menasria and S. Miloudi for excellent technical assistance. The authors acknowledge the work of technicians and engineers of the Biology and Pathology Center of the Lille University Hospital (CHU Lille) who are involved in the diagnostics of SARS-CoV-2. The authors acknowledge the contribution of the Lille COVID Research Network (LICORNE).

## Ethical approval

The study was performed in accordance with the Declaration of Helsinki. The study was approved by the French Institutional Authority for Personal Data Protection (Commission Nationale de l’Informatique et des Libertés DR-2020-178, October 22nd, 2020) and the ethics committee (Comité de Protection des Personnes Nord Ouest IV, ECH20/09, September 7th, 2020).

## Conflicts of Interest

The authors declare no conflict of interest.

## Funding

This research was supported by I-SITE ULNE, the Centre Hospitalier Universitaire de Lille and Université de Lille.

## Notes

### Competing Interest Statement

The authors have declared no competing interest.

### Funding Statement

This research was supported by I-SITE ULNE, the Centre Hospitalier Universitaire de Lille and University of Lille.

### Author Declarations

The study was performed in accordance with the Declaration of Helsinki. The study was approved by the French Institutional Authority for Personal Data Protection (Commission Nationale de l Informatique et des Libertes DR-2020-178, October 22nd, 2020) and the ethics committee (Comite de Protection des Personnes Nord Ouest IV, ECH20/09, September 7th, 2020).

