## Supplementary Tables for "Altered microRNA expression in severe COVID-19: potential prognostic and pathophysiological role"

### Supplementary Table 1. Patients' characteristics

| | COVID-19 | | Control |
|---|---|---|---|
| | Non severe | Severe | |
| Number of cases | 21 | 20 | 20 |
| M/F | 10/11 | 14/6 | 9/11 |
| Age (median (range)) | 30 (23 – 62) | 60 (29 – 78) | 71 (23 – 89) |
| Days post symptom onset (median) | 7 | 7 | NA |
| Hospital admission, n (%) | 0 | 20 (100%) | 19 (95%) |
| **Biologic tests;** median (n with available data) | | | |
| Hemoglobin (g/dL) | NA | 13.5 (n=20) | 12.4 (n=18) |
| Platelets ($10^9$/L) | NA | 197 (n=20) | 220 (n=18) |
| Leukocytes ($10^9$/L) | NA | 6.2 (n=20) | 9.8 (n=18) |
| CRP (mg/L) | NA | 93 (n=20) | 89 (n=19) |
| LDH (U/L) | NA | 449 (n=15) | 262 (n=5) |
| **ICU characteristics** | | | |
| Intensive care unit stay, n (%) | 0 | 20 (100%) | 5 (25%) |
| Oxygen treatment, n (%) | 0 | 20 (100%) | 7 (35%) |
| SOFA[†] score at admission (median) | NA | 6.5 | 2 |
| Score WHO[‡] at admission (median) | NA | 6 | NA |
| Score WHO[‡] day 28 (median) | NA | 4 (n=16) | NA |
| Oxygen by mask high concentration, n (%) | NA | 9 (45%) | 1 (2.5%) |
| Optiflow, n (%) | NA | 13 (65%) | 1 (2.5%) |
| Non-invasive ventilation, n (%) | NA | 9 (45%) | 2 (5%) |
| Invasive mechanical ventilation, n (%) | NA | 16 (80%) | 2 (5%) |
| Extracorporeal membrane oxygenation, n (%) | NA | 1 (5%) | 0 |
| **Underlying diseases**, n (%) | | | |
| Kidney disease | 0 | 1 (5%) | 3 (15%) |
| Hypertension | 3 (14%) | 9 (45%) | 9 (45%) |
| Cardiovascular diseases | 2 (10%) | 3 (15%) | 8 (40%) |
| Respiratory system disease | 3 (14%) | 5 (25%) | 6 (30%) |
| Diabetes | 0 | 3 (15%) | 6 (30%) |
| Cancer | 0 | 0 | 2 (10%) |
| Malignant haemopathy | 0 | 1 (5%) | 1 (5%) |
| Immunosuppression | 1 (5%) | 1 (5%) | 3 (15%) |

[†]SOFA: Sequential Organ Failure Assessment Score

[‡]WHO: world health organization

### Supplementary Table 2. Target genes

| Entrezid | Genesymbol | Description |
|---|---|---|
| 6310 | ATXN1 | Homo sapiens ataxin 1 (ATXN1), transcript variant 1, mRNA. |
| 596 | BCL2 | Homo sapiens BCL2 apoptosis regulator (BCL2), transcript variantalpha, mRNA. |
| 490 | ATP2B1 | Homo sapiens ATPase plasma membrane Ca2+ transporting 1 (ATP2B1),transcript variant 1, mRNA. |
| 59338 | PLEKHA1 | Homo sapiens pleckstrin homology domain containing A1 (PLEKHA1),transcript variant 2, mRNA. |
| 23131 | GPATCH8 | Homo sapiens G-patch domain containing 8 (GPATCH8), transcriptvariant 1, mRNA. |
| 51114 | ZDHHC9 | Homo sapiens zinc finger DHHC-type palmitoyltransferase 9 (ZDHHC9),transcript variant 2, mRNA. |
| 605 | BCL7A | Homo sapiens BAF chromatin remodeling complex subunit BCL7A(BCL7A), transcript variant 2, mRNA. |
| 163 | AP2B1 | Homo sapiens adaptor related protein complex 2 subunit beta 1(AP2B1), transcript variant 1, mRNA. |
| 91746 | YTHDC1 | Homo sapiens YTH domain containing 1 (YTHDC1), transcript variant1, mRNA. |
| 6812 | STXBP1 | Homo sapiens syntaxin binding protein 1 (STXBP1), transcriptvariant 2, mRNA. |
| 7335 | UBE2V1 | Homo sapiens ubiquitin conjugating enzyme E2 V1 (UBE2V1),transcript variant 4, mRNA. |
| 221937 | FOXK1 | Homo sapiens forkhead box K1 (FOXK1), mRNA. |
| 56990 | CDC42SE2 | Homo sapiens CDC42 small effector 2 (CDC42SE2), transcript variant2, mRNA. |
| 3955 | LFNG | Homo sapiens LFNG O-fucosylpeptide3-beta-N-acetylglucosaminyltransferase (LFNG), transcript variant |
| 440193 | CCDC88C | Homo sapiens coiled-coil domain containing 88C (CCDC88C), mRNA. |
| 25994 | HIGD1A | Homo sapiens HIG1 hypoxia inducible domain family member 1A(HIGD1A), transcript variant 1, mRNA; nuclear gene for |
| 90809 | PIP4P1 | Homo sapiens phosphatidylinositol-4,5-bisphosphate 4-phosphatase 1(PIP4P1), transcript variant 1, mRNA. |
| 7763 | ZFAND5 | Homo sapiens zinc finger AN1-type containing 5 (ZFAND5), transcriptvariant a, mRNA. |
| 996 | CDC27 | Homo sapiens cell division cycle 27 (CDC27), transcript variant 1,mRNA. |
| 1111 | CHEK1 | Homo sapiens checkpoint kinase 1 (CHEK1), transcript variant 2,mRNA |
| 5255 | PHKA1 | Homo sapiens phosphorylase kinase regulatory subunit alpha 1(PHKA1), transcript variant 2, mRNA. |
| 3977 | LIFR | Homo sapiens LIF receptor subunit alpha (LIFR), transcript variant1, mRNA. |
| 813 | CALU | Homo sapiens calumenin (CALU), transcript variant 2, mRNA. |
| 9209 | LRRFIP2 | Homo sapiens LRR binding FLII interacting protein 2 (LRRFIP2),transcript variant 3, mRNA. |
| 64225 | ATL2 | Homo sapiens atlastin GTPase 2 (ATL2), transcript variant 2, mRNA. |
| 27 | ABL2 | Homo sapiens ABL proto-oncogene 2, non-receptor tyrosine kinase(ABL2), transcript variant d, mRNA. |

| | | |
|---|---|---|
| 896 | CCND3 | Homo sapiens cyclin D3 (CCND3), transcript variant 1, mRNA. |
| 552889 | ATXN7L3B | Homo sapiens ataxin 7 like 3B (ATXN7L3B), mRNA. |
| 2113 | ETS1 | Homo sapiens ETS proto-oncogene 1, transcription factor (ETS1),transcript variant 1, mRNA. |
| 7465 | WEE1 | Homo sapiens WEE1 G2 checkpoint kinase (WEE1), transcript variant2, mRNA. |
| 8867 | SYNJ1 | Homo sapiens synaptojanin 1 (SYNJ1), transcript variant 3, mRNA. |
| 651746 | ANKRD33B | Homo sapiens ankyrin repeat domain 33B (ANKRD33B), mRNA. |
| 11228 | RASSF8 | Homo sapiens Ras association domain family member 8 (RASSF8),transcript variant 1, mRNA. |
| 55206 | SBNO1 | Homo sapiens strawberry notch homolog 1 (SBNO1), transcript variant1, mRNA. |
| 63967 | CLSPN | Homo sapiens claspin (CLSPN), transcript variant 2, mRNA. |
| 4781 | NFIB | Homo sapiens nuclear factor I B (NFIB), transcript variant 1, mRNA. |
| 79751 | SLC25A22 | Homo sapiens solute carrier family 25 member 22 (SLC25A22),transcript variant 1, mRNA; nuclear gene for mitochondrial product. |
| 51444 | RNF138 | Homo sapiens ring finger protein 138 (RNF138), transcript variant3, mRNA. |
| 7049 | TGFBR3 | Homo sapiens transforming growth factor beta receptor 3 (TGFBR3),transcript variant 2, mRNA. |
| 862 | RUNX1T1 | Homo sapiens RUNX1 partner transcriptional co-repressor 1(RUNX1T1), transcript variant 5, mRNA. |
| 5451 | POU2F1 | Homo sapiens POU class 2 homeobox 1 (POU2F1), transcript variant 2,mRNA. |
| 83607 | AMMECR1L | Homo sapiens AMMECR1 like (AMMECR1L), transcript variant 2, mRNA. |
| 57154 | SMURF1 | Homo sapiens SMAD specific E3 ubiquitin protein ligase 1 (SMURF1),transcript variant 3, mRNA. |
| 9354 | UBE4A | Homo sapiens ubiquitination factor E4A (UBE4A), transcript variant2, mRNA. |
| 10000 | AKT3 | Homo sapiens AKT serine/threonine kinase 3 (AKT3), transcriptvariant 3, mRNA. |
| 905 | CCNT2 | Homo sapiens cyclin T2 (CCNT2), transcript variant a, mRNA. |
| 5110 | PCMT1 | Homo sapiens protein-L-isoaspartate (D-aspartate)O-methyltransferase (PCMT1), transcript variant 2, mRNA. |
| 55334 | SLC39A9 | Homo sapiens solute carrier family 39 member 9 (SLC39A9),transcript variant 2, mRNA. |
| 10802 | SEC24A | Homo sapiens SEC24 homolog A, COPII coat complex component(SEC24A), transcript variant 2, mRNA. |
| 390 | RND3 | Homo sapiens Rho family GTPase 3 (RND3), transcript variant 1,mRNA. |
| 8945 | BTRC | Homo sapiens beta-transducin repeat containing E3 ubiquitin proteinligase (BTRC), transcript variant 3, mRNA. |
| 92 | ACVR2A | Homo sapiens activin A receptor type 2A (ACVR2A), transcriptvariant 1, mRNA. |
| 10558 | SPTLC1 | Homo sapiens serine palmitoyltransferase long chain base subunit 1(SPTLC1), transcript variant 3, mRNA. |
| 7227 | TRPS1 | Homo sapiens transcriptional repressor GATA binding 1 (TRPS1),transcript variant 2, mRNA. |

| | | |
|---|---|---|
| 10152 | ABI2 | Homo sapiens abl interactor 2 (ABI2), transcript variant 1, mRNA. |
| 23271 | CAMSAP2 | Homo sapiens calmodulin regulated spectrin associated proteinfamily member 2 (CAMSAP2), transcript variant 1, mRNA. |
| 9444 | QKI | Homo sapiens QKI, KH domain containing RNA binding (QKI),transcript variant 5, mRNA. |
| 51621 | KLF13 | Homo sapiens Kruppel like factor 13 (KLF13), transcript variant 2,mRNA. |
| 22880 | MORC2 | Homo sapiens MORC family CW-type zinc finger 2 (MORC2), transcriptvariant 1, mRNA. |
| 23471 | TRAM1 | Homo sapiens translocation associated membrane protein 1 (TRAM1),transcript variant 2, mRNA. |
| 1788 | DNMT3A | Homo sapiens DNA methyltransferase 3 alpha (DNMT3A), transcriptvariant 5, mRNA. |
| 80851 | SH3BP5L | Homo sapiens SH3 binding domain protein 5 like (SH3BP5L),transcript variant 2, mRNA. |
| 57089 | ENTPD7 | Homo sapiens ectonucleoside triphosphate diphosphohydrolase 7(ENTPD7), transcript variant 1, mRNA. |
| 6774 | STAT3 | Homo sapiens signal transducer and activator of transcription 3(STAT3), transcript variant 4, mRNA. |
| 23030 | KDM4B | Homo sapiens lysine demethylase 4B (KDM4B), transcript variant 2,mRNA. |
| 64393 | ZMAT3 | Homo sapiens zinc finger matrin-type 3 (ZMAT3), transcript variant3, mRNA. |
| 2683 | B4GALT1 | Homo sapiens beta-1,4-galactosyltransferase 1 (B4GALT1), transcriptvariant 2, mRNA. |
| 1951 | CELSR3 | Homo sapiens cadherin EGF LAG seven-pass G-type receptor 3(CELSR3), mRNA. |
| 1810 | DR1 | Homo sapiens down-regulator of transcription 1 (DR1), mRNA. |
| 5713 | PSMD7 | Homo sapiens proteasome 26S subunit, non-ATPase 7 (PSMD7), mRNA. |
| 6664 | SOX11 | Homo sapiens SRY-box transcription factor 11 (SOX11), mRNA. |
| 8760 | CDS2 | Homo sapiens CDP-diacylglycerol synthase 2 (CDS2), mRNA. |
| 6197 | RPS6KA3 | Homo sapiens ribosomal protein S6 kinase A3 (RPS6KA3), mRNA. |
| 7189 | TRAF6 | Homo sapiens TNF receptor associated factor 6 (TRAF6), transcriptvariant 2, mRNA. |
| 9371 | KIF3B | Homo sapiens kinesin family member 3B (KIF3B), mRNA. |
| 1795 | DOCK3 | Homo sapiens dedicator of cytokinesis 3 (DOCK3), mRNA. |
| 1650 | DDOST | Homo sapiens dolichyl-diphosphooligosaccharide--proteinglycosyltransferase non-catalytic subunit (DDOST), mRNA. |
| 1820 | ARID3A | Homo sapiens AT-rich interaction domain 3A (ARID3A), mRNA. |
| 830 | CAPZA2 | Homo sapiens capping actin protein of muscle Z-line subunit alpha 2(CAPZA2), mRNA. |
| 10527 | IPO7 | Homo sapiens importin 7 (IPO7), mRNA. |
| 7832 | BTG2 | Homo sapiens BTG anti-proliferation factor 2 (BTG2), mRNA. |
| 5930 | RBBP6 | Homo sapiens RB binding protein 6, ubiquitin ligase (RBBP6),transcript variant 1, mRNA. |

| | | |
|---|---|---|
| 6515 | SLC2A3 | Homo sapiens solute carrier family 2 member 3 (SLC2A3), mRNA. |
| 23011 | RAB21 | Homo sapiens RAB21, member RAS oncogene family (RAB21), mRNA. |
| 51592 | TRIM33 | Homo sapiens tripartite motif containing 33 (TRIM33), transcriptvariant a, mRNA. |
| 51132 | RLIM | Homo sapiens ring finger protein, LIM domain interacting (RLIM),transcript variant 1, mRNA. |
| 54887 | UHRF1BP1 | Homo sapiens UHRF1 binding protein 1 (UHRF1BP1), mRNA. |
| 55664 | CDC37L1 | Homo sapiens cell division cycle 37 like 1 (CDC37L1), mRNA. |
| 55824 | PAG1 | Homo sapiens phosphoprotein membrane anchor with glycosphingolipidmicrodomains 1 (PAG1), mRNA. |
| 56977 | STOX2 | Homo sapiens storkhead box 2 (STOX2), transcript variant 1, mRNA. |
| 57534 | MIB1 | Homo sapiens mindbomb E3 ubiquitin protein ligase 1 (MIB1), mRNA. |
| 57551 | TAOK1 | Homo sapiens TAO kinase 1 (TAOK1), transcript variant 1, mRNA. |
| 53354 | PANK1 | Homo sapiens pantothenate kinase 1 (PANK1), transcript variantgamma, mRNA. |
| 134353 | LSM11 | Homo sapiens LSM11, U7 small nuclear RNA associated (LSM11), mRNA. |
| 387522 | PEDS1-UBE2V1 | Homo sapiens PEDS1-UBE2V1 readthrough (PEDS1-UBE2V1), mRNA. |

### Supplementary Table 3. GO biological processes

| Name | Pvalue | adjusted Pvalue(BH) |
|---|---|---|
| GO:0000209_protein_polyubiquitination | <0.001 | <0.001 |
| GO:0006974_cellular_response_to_DNA_damage_stimulus | <0.001 | <0.001 |
| GO:0019886_antigen_processing_and_presentation_of_exogenous_peptide_antigen_via_MHC_class_II | <0.001 | 0.001 |
| GO:0006511_ubiquitin-dependent_protein_catabolic_process | 0.006 | 0.014 |
| GO:0035556_intracellular_signal_transduction | 0.019 | 0.037 |

### Supplementary Table 4. GO molecular functions

| Name | Pvalue | adjusted Pvalue(BH) |
|---|---|---|
| GO:0004672_protein_kinase_activity | <0.001 | 0.002 |
| GO:0030165_PDZ_domain_binding | <0.001 | 0.002 |
| GO:0004842_ubiquitin-protein_transferase_activity | <0.001 | 0.002 |
| GO:0061630_ubiquitin_protein_ligase_activity | 0.002 | 0.005 |
| GO:0019901_protein_kinase_binding | 0.006 | 0.011 |
| GO:0000978_RNA_polymerase_II_proximal_promoter_sequence-specific_DNA_binding | 0.026 | 0.040 |
| GO:0001228_DNA-binding_transcription_activator_activity_RNA_polymerase_II-specific | 0.043 | 0.048 |
| GO:0008134_transcription_factor_binding | 0.042 | 0.048 |
